# Summer COVID-19 third wave: faster high altitude spread suggests high UV adaptation

**DOI:** 10.1101/2020.08.17.20176628

**Authors:** Hervé Seligmann, Nicolas Vuillerme, Jacques Demongeot

## Abstract

We present spread parameters for first and second waves of the COVID-19 pandemy for USA states, and third wave for 32 regions (19 countries and 13 states of the USA) detected beginning of August 2020. USA first/second wave spreads increase/decrease with population density, are uncorrelated with temperature and median population age. Pooling all 32 regions, third wave spread is slower than for first wave, similar to second wave, and increases with mean altitude (second wave slopes decrease above 900m). Apparently, viruses adapted in spring (second wave) to high temperatures and infecting the young, and in summer (third) waves for spread at altitudes above 1000m. Third wave slopes are not correlated to temperature, so patterns with elevation presumably indicate resistance to relatively high UV regimes. Environmental trends of the COVID-19 pandemy change at incredible rates, making predictions based on classical epidemiological knowledge particularly uncertain.

## Introduction

Spread parameters of daily new confirmed COVID-19 cases estimate viral contagiousness. Previously, it was shown that first wave spread, when comparing different countries, decreases with mean annual temperature (Demongeot et al 2020). The opposite trend with temperature occurs for second wave spread parameters (Seligmann et al 2020).

This is in line with observations on variation in spread across different regions of Italy, in March 2020 (first wave period, negative correlation with temperature) and in May 2020 (second wave period, positive correlation with temperature) (Malki et al 2020).

First and second wave spreads differ also in terms of other factors: first/second wave spread increases/decreases with median population age, and decreases/increases later their respective onset dates. First and second waves also differ because we detected no associations between second wave slopes and mean country elevation, while first wave slopes increase with elevation up to 900m, and decrease beyond that approximate altitude (Seligmann et al 2020).

Inversion of trends for these independent covariates is difficult to explain. One could invoke different explanations for each factor, but a common, most parsimonious explanation would involve deterministic mutation dynamics, resulting in parallel evolution of distinct virus populations (Forster et al 2020). Our hypothesis is that self-hybridization of the virus’ singlestranded RNA genomes protects nucleotides forming stems against mutations, while favouring mutations in loops. Mutation cumulation presumably cause deterministic switches between few optimal structures which differ in their properties in relation to temperature, etc. Such switches between secondary structures have been suggested for COVID-19 after small insertion/deletion mutations (Manzourolajdad et al 2020).

These patterns remind the little-known phenomenon of negative heritability. Usually, heritabilities of traits are such that offsprings resemble their parents: if a parent is for a given trait above average, on average, his/her offspring will also be above average, and parents below average for that trait have on average offspring which are below average for that trait. In short, heritabilities are correlations between traits of parents and offspring. These correlations are usually positive. Surprisingly, in some cases, negative correlations were observed, meaning negative heritabilities, where above average parents produce below average offspring for given traits, and vice versa for below average parents (Janssen et al 1988, Haldane 1996). These phenomena are not statistical artefacts (Steinsaltz et al 2020), but are difficult to reproduce empirically (Stam et al 2017). This is expected when assuming negative heritability results from selection under changing environmental conditions (Bonduriansky et al 2011).

Hence, according to the principle of trend inversion under changing environments, we expect that spread parameters for the third wave correlate as observed for the first wave. We also report first and second wave parameters for the 50 states of the USA, and describe their environmental covariates.

## Materials and methods

We used the methods as in previous analyses (Demongeot et al 2020, Seligmann et al 2020). The coefficients (slopes) of regression analyses are considered as estimates of viral spread. We adjust the exponential model y = a*exp(b*x), where y is the daily number of new confirmed COVID-19 cases, x is the number of days since wave onset, a is a constant and b is the slope. The log-transformed version of this is ln y = ln a + b*x. Daily numbers of new cases and total numbers of tests per countries are from the site www.worldometers.info/coronavirus/ (accessed on 31 July 2020).

Determination of first wave onset is defined by the date when the cumulative number of cases is closest to 100 cases. Second and third waves are determined visually by eyeballing graphs plotting daily new confirmed cases as a function of days since February 15, 2020.

For countries, sources for mean elevation, mean temperature, population density and median population age are as previously (Seligmann et al 2020). For the 50 states of the USA, we used the following sources:

**Table 1.** Web sites for data sources

|  |  |
| --- | --- |
| temperature | https://www.currentresults.com/Weather/US/average-annual-state-temperatures.php |
| elevation | https://en.wikipedia.org/wiki/List_of_U.S._states_and_territories_by_elevation |
| density | https://en.wikipedia.org/wiki/List_of_states_and_territories_of_the_United_States_by_population_density |
| median age | https://en.wikipedia.org/wiki/List_of_U.S._states_and_territories_by_median_age. |

## Results

### First, second and third wave for Wisconsin

Figure 1 plots daily numbers of confirmed new cases in Wisconsin, USA. Visual inspection of Figure 1 detects a second wave from 20/4 until end of May, and a third wave from 14/6 until end of July. The highest slope is for the first wave, second and third wave slopes are about a third of the first wave slope.

**Figure 1.**
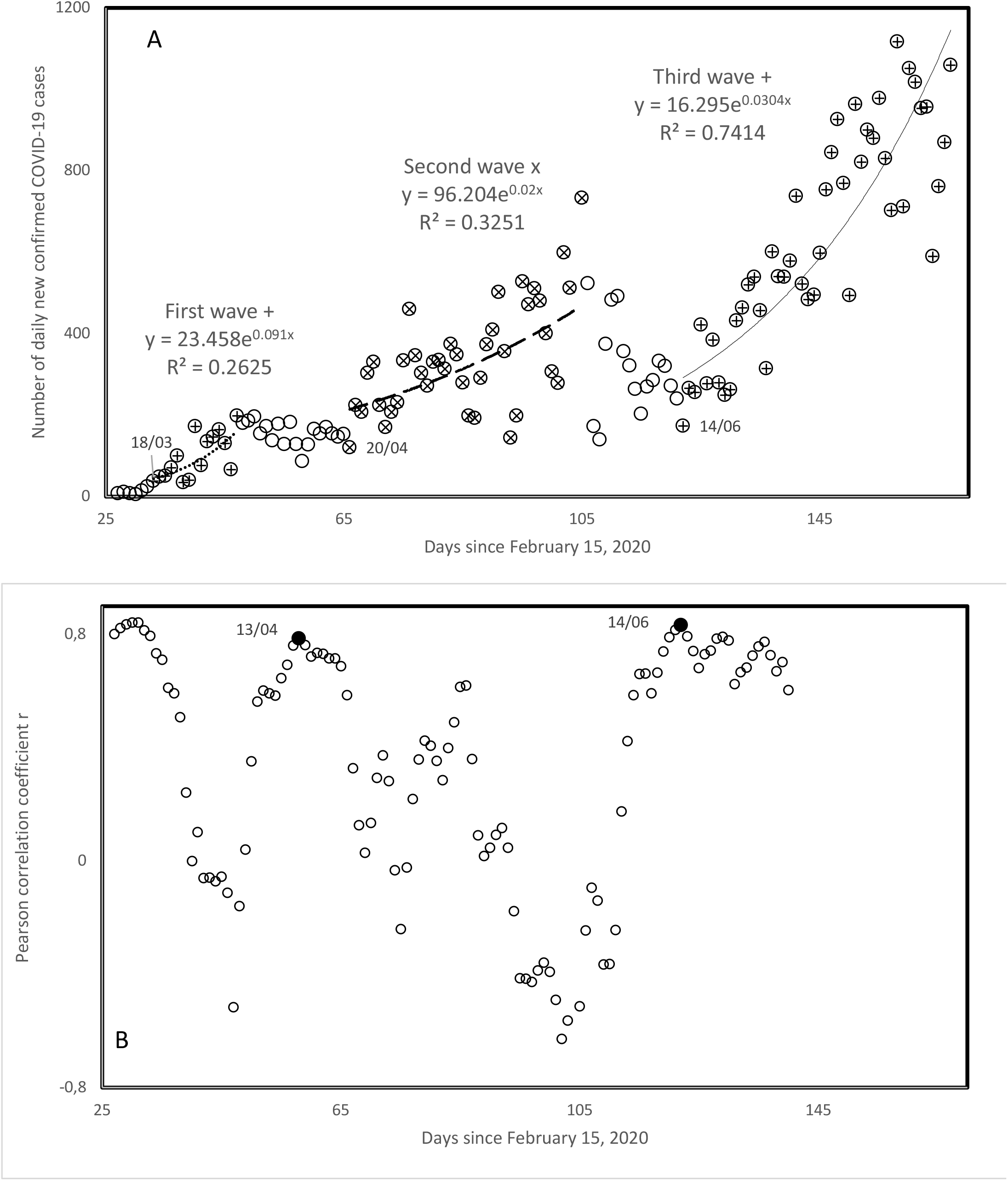
A. Daily confirmed COVID-19 cases in Wisconsin, as a function of days since Februrary 15. Visual inspection determined second/third waves. B. Pearson correlation coefficient r calculated for a running window of 20 consecutive days in A, as a function of days since Februrary 15 for the first day included in the running window. Second/third wave onsets (filled circles) are determined by local maxima in r.

### Spread parameters for the USA

Table 2 presents for each of the 50 states of the USA onset dates and slopes of all three waves, as detected by visual inspection of figures as Figure 1, for all remaining states. At mid-August, 42 among 50 states have a second wave. Second wave slopes are lower than first wave slope in all but three states, Oklahoma (slopes basically identical), and Kansas and Ohio (second wave spread greater than first wave spread). Hence, overall, spread decreased from the first to the second wave, from mean first wave slope = 0.1902 to mean second wave slope = 0.0585.

**Table 2.**
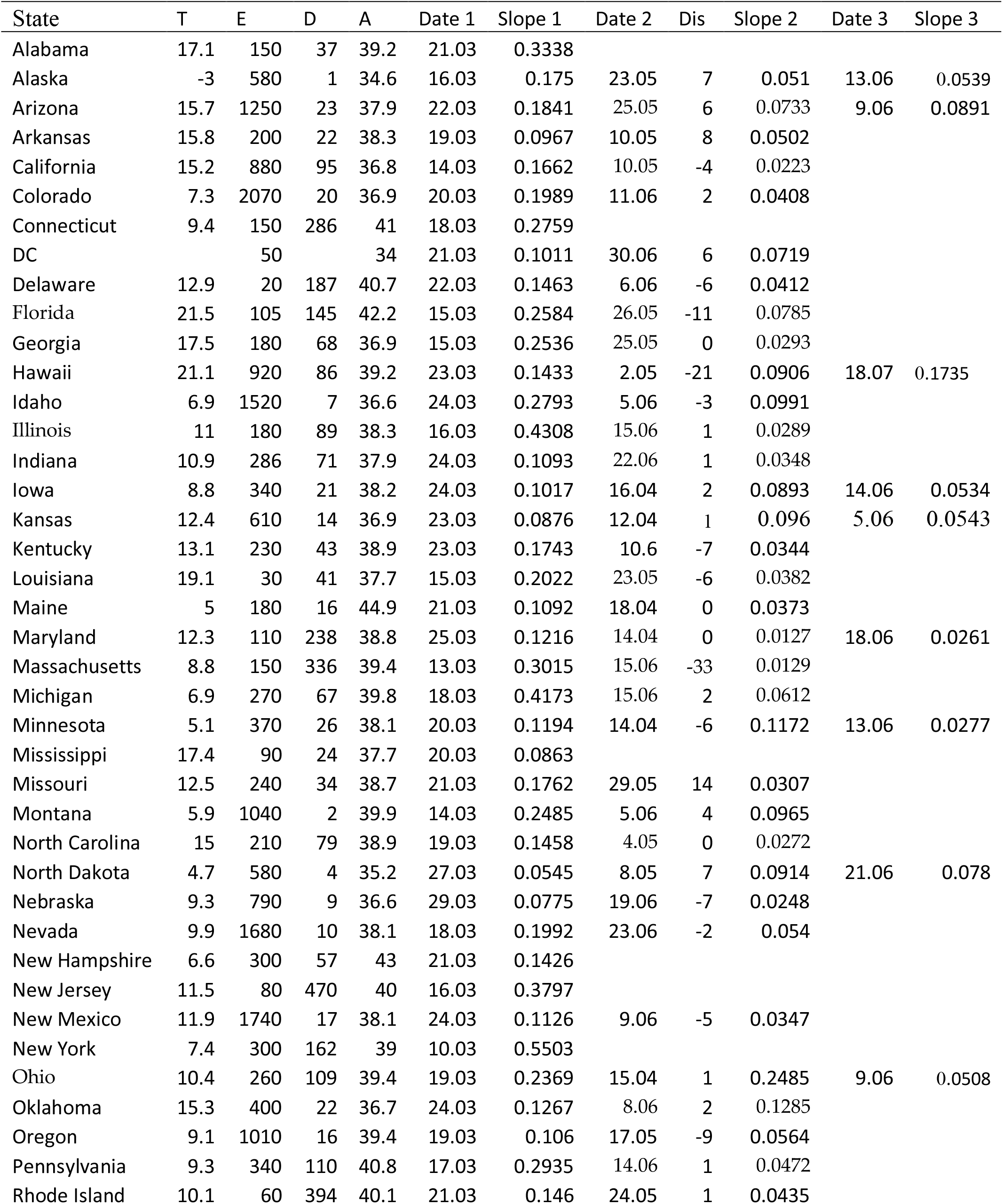
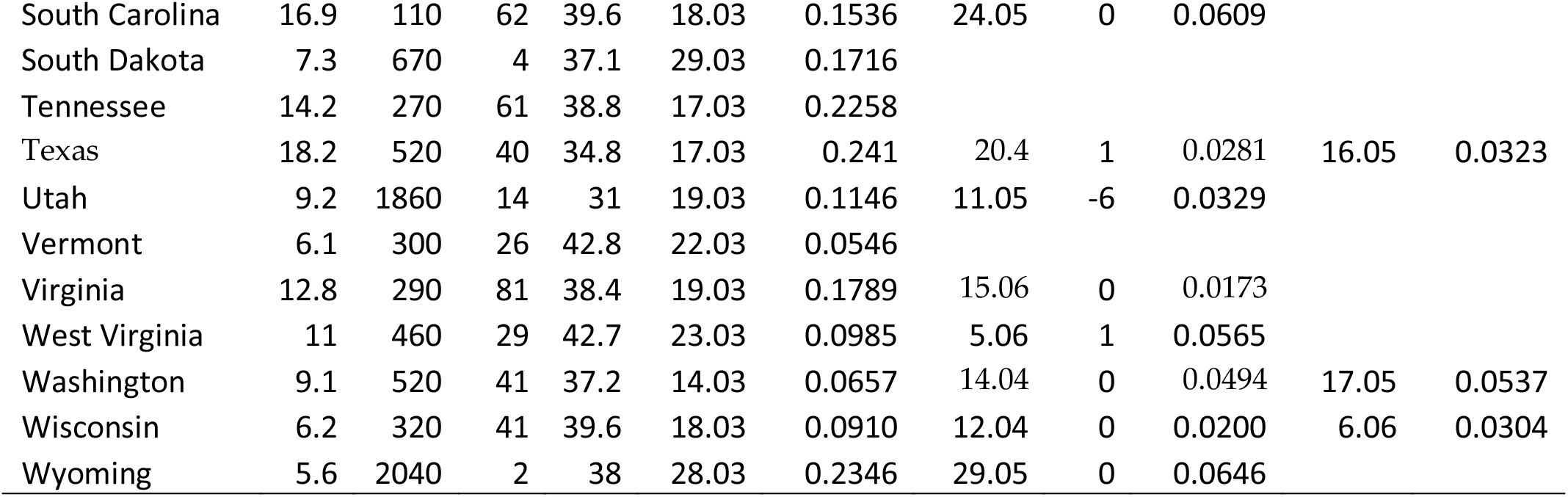
Spread parameters of first, second and third waves in the 50 states of the USA. Columns are: 1. State, 2. Mean annual temperature, 3. Mean elevation, 4. Population density, 5. Median age, 6. Onset date of first wave, 7. Exponential slope of first wave, 8. Onset date of second wave, 9. Days separating onset date in column 8 vs onset date determined by statistical analysis as in Figure 1 B, 10. Exponential slope of second wave, 11. Onset date of third wave, 12. Exponential slope of third wave.

There are third waves at mid-August in 13 states, mean slope = 0.0602. Third wave slopes are weaker than first wave slopes in eleven among thirteen cases, besides in Hawaii and North Dakota, where the third wave spread is faster than the first wave spread. As compared to the second wave, third wave slopes are greater in six states. Hence, second and third waves spread rates are similar.

In the USA, first wave slopes decrease with time since first wave onset (r = −0.5306, two-tailed P = 0.000074). This is similar to the decrease in slopes previously reported for comparisons across countries (Seligmann et al 2020). Comparing countries, second wave slopes increase with time since second wave onset (Seligmann et al 2020). However, for the USA, no such increase could be detected, the trend might fit with that observed for the first wave (r = −0.29, P = 0.06). Also contrasting with previous analyses of variation between countries, there is no association in the USA between first and second wave slopes and mean annual temperature.

A positive association exists between first wave slope and time since second wave onset (r = 0.4469, two-tailed P = 0.003), which could indicate that high initial spread rates contribute to early onset of ulterior waves. Note that this effect does not explain the absence of second waves in several states with particularly high first waves, such as Illinois, New Jersey, New York.

First wave slopes increase with population density (r = 0.3777, one-tailed P = 0.0034, Figure 2A). This association is expected, but was not observed when comparing different countries (Demongeot et al 2020, Seligmann et al 2020). Six New England states have relatively low slopes considering their densities. Excluding them from calculations increases the strength of this correlation with density (r = 0.5538, one-tailed P = 0.00005).

**Figure 2.**
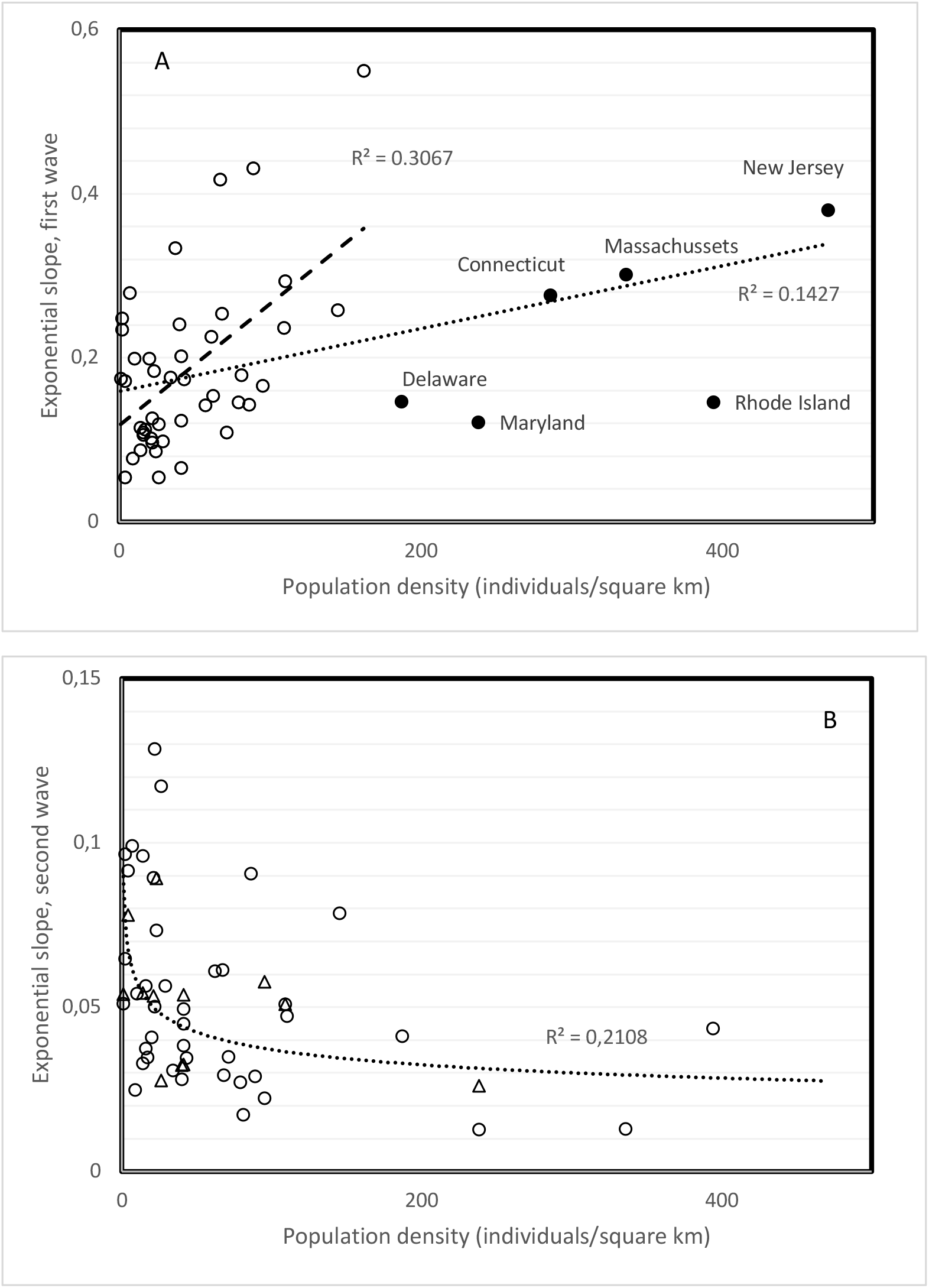
Spread parameters (slope of exponential regression) of first (A) and second (B) waves in states of the USA as a function of population density. Trend inverted between first and second wave. A: the New England states with high densities (filled circles) follow a different pattern than the rest of states, with a slower increase in spread with increasing density. B: the second wave slopes are overall lower than first wave slopes. Third wave slopes (filled triangles) follow second wave trend with population density.

Density in principle increases spread rate. This was not observed for first and second wave slopes when comparing countries (Demongeot et al 2020, Seligmann et al 2020). However, this density principle is confirmed for first wave slopes from the USA (Figure 2A). Unexpectedly, second wave slopes decrease with population density (r = −0.459, two-tailed P = 0.0022, Figure 2B). This unexplained inversion of patterns between first and second waves has been reported for covariates such as temperature, time since wave onset and median population age when comparing different countries (Seligmann et al 2020). The decrease of slopes with density in the USA is foremost astounding because it does not at all fit with any knowledge on disease spread.

We find two associations between third wave variables for the USA and other variables. The third wave slope increases with time since second wave onset (r = 0.5749, two tailed P = 0.0398). Third wave slope also increases with mean altitude (r = 0.731, two tailed P = 0.0045). Only the latter association was confirmed by analyses of variation among countries below.

### Visual inspection vs objective statistical analysis

One can argue that determining onsets of waves from visual examinations of graphs is subjective. This issue has been raised in the past (Lund and Reeves 2002, Percival and Rothrock 2005), but has no obvious simple solutions and requires extensive simulation analyses curtailed to each specific dataset, meaning in this case for each state (Ghorbanzadh and Picard 1992, Ondo 2002). We therefore use a simplified method. Statistical analyses based on calculating Pearson correlation coefficients r for a running window of 20 days determine a local maximum for r within five days of the second wave onset date determined visually for 80 percent of the countries examined (Seligmann et al 2020). Figure 1B plots these r values for Wisconsin as a function of the first day included in the running window. Onset dates are close to those indicated in Figure 1A. We applied this method for the 42 second waves detected for the USA (Dis, Table 2). Most of the onset dates determined by running windows (62%) are within five days of the date determined visually, in line with similar previous analyses for other countries (Seligmann et al 2020).

### Third waves in 19 countries

Among 123 countries for which we examined the daily number of new confirmed cases as a function of time, we found second waves in 73 countries, including 28 countries for which the second wave was previously reported (Seligmann et al 2020). Among these, by mid-August, there were third waves in 19 countries (Table 3). Second wave slopes are lower than first waves in the vast majority of countries with 17 exceptions (23 %), third wave slopes are lower than first wave slopes in five countries, and lower than second wave slopes in about half the countries.

**Table 3.** Spread parameters of first, second and third waves in countries across the world. This table includes all countries for which third waves were detected, and those for which second waves were not included in Seligmann et al (2020) because undetected at that time. Columns are as in Table 2. Countries with missing first wave estimates are due to sporadic data or a fast increase spanning over too few days for valid statistical estimation.

| Country | T | E | D | A | Date<br>1 | Slope 1 | Date 2 | Slope 2 | Date 3 | Slope 3 |
| --- | --- | --- | --- | --- | --- | --- | --- | --- | --- | --- |
| Africa |  |  |  |  |  |  |  |  |  |  |
| Algeria | 22.5 | 800 | 18 | 28.1 | 20.03 | 0.1594 | 7.04 | 0.0316 | 4.06 | 0.0369 |
| Djibouti | 28.0 | 430 | 47 | 23.9 | 7.04 | 0.489 |  |  |  |  |
| Guinea | 25.7 | 472 | 50 | 18.9 | 4.04 | 0.0744 |  |  |  |  |
| Mauritius | 22.4 | 209 | 620 | 35.3 | 28.03 | 0.0972 |  |  |  |  |
| Morocco | 17.1 | 909 | 80 | 29.3 | 21.03 | 0.1161 | 31.05 | 0.0687 | 21.07 | 0.0894 |
| Nigeria | 26.8 | 30 | 218 | 18.4 | 29.03 | 0.0672 | 2.05 | 0.0258 |  |  |
| Senegal | 27.85 | 69 | 82 | 18.8 |  |  | 12.04 | 0.1003 | 17.05 | 0.0238 |
| Sudan | 26.9 | 568 | 22 | 19.9 | 20.04 | 0.1032 |  |  |  |  |
| Tunisia | 19.2 | 246 | 186 | 31.6 |  |  | 24.06 | 0.1422 |  |  |
| Asia |  |  |  |  |  |  |  |  |  |  |
| Bangladesh | 25.0 | 85 | 1175 | 26.7 | 5.04 | 0.2409 |  |  |  |  |
| Iran | 17.25 |  | 51 | 30.3 | 26.02 | 0.2641 | 1.05 | 0.0438 | 26.05 | 0.0666 |
| Iraq | 14.03 | 312 | 90 | 20.0 | 14.03 | 0.1184 | 14.03 | 0.041 | 17.07 | 0.0287 |
| Israel | 19.2 | 508 | 417 | 29.9 |  |  | 24.05 | 0.1071 |  |  |
| Japan | 11.15 | 438 | 333 | 47.3 | 20.02 | 0.0872 | 24.05 | 0.0381 | 26.06 | 0.1132 |
| Jordan | 18.3 | 812 | 120 | 22.5 | 18.03 | 0.1421 | 28.04 | 0.2977 | 25.05 | 0.1034 |
| Kazakhstan | 6.4 | 387 | 7 | 30.6 | 26.03 | 0.0856 | 8.05 | 0.0933 | 6.06 | 0.0636 |
| Kuwait | 25.35 | 108 | 642 | 29.3 | 12.03 | 0.1031 | 14.06 | 0.0512 |  |  |
| Kyrgyzstan | 1.55 | 2988 | 32 | 26.5 | 30.03 | 0.0671 | 25.04 | 0.0274 | 2.06 | 0.104 |
| Nepal | 8.1 | 3265 | 201 | 24.1 | 06.05 | 0.207 | 18.07 | 0.0729 |  |  |
| Pakistan | 20.20 | 900 | 274 | 23.8 | 15.03 | 0.1301 | 27.05 | 0.0696 |  |  |
| Qatar | 27.15 | 28 | 237 | 33.2 |  |  | 2.08 | 0.0626 |  |  |
| Turkey | 11.1 | 1132 | 106 | 30.9 |  |  | 2.06 | 0.0473 |  |  |
| Europa |  |  |  |  |  |  |  |  |  |  |
| Albania | 11.4 | 708 | 100 | 32.9 | 23.03 | 0.0309 | 18.05 | 0.0825 | 1.06 | 0.0991 |
| Austria | 6.35 | 910 | 106 | 44.0 | 08/3 | 0.2825 | 7.06 | 0.0545 |  |  |
| Belgium | 9.55 | 181 | 378 | 41.4 | 06/3 | 0.1963 | 16.06 | 0.0257 |  |  |
| Bosnia/Her. | 9.85 | 500 | 69 | 42.1 | 21.03 | 0.1671 | 5.06 | 0.0667 |  |  |
| Bulgaria | 10.55 | 472 | 64 | 42.7 | 19.03 | 0.0927 | 13.04 | 0.1163 | 25.05 | 0.1255 |
| Croatia | 10.9 | 331 | 187 | 43 | 18.03 | 0.093 | 12.06 | 0.3213 |  |  |
| Denmark | 7.5 | 34 | 349 | 42.2 |  |  | 7.07 | 0.0539 |  |  |
| France | 10.7 | 375 | 123 | 41.4 | 29.02 | 0.2898 | 24.06 | 0.0308 |  |  |
| Finland | 1.7 | 134 | 16 | 42.5 | 12.03 | 0.0711 | 26.07 | 0.0891 |  |  |
| Greece | 15.4 | 498 | 210 | 44.5 | 11.03 | 0.0759 | 24.06 | 0.0854 |  |  |
| Georgia | 5.8 | 1432 | 54 | 38.1 | 30.03 | 0.0346 | 2.06 | 0.1471 | 22.06 | 0.0481 |
| Germany | 8.5 | 263 | 233 | 47.1 | 29.02 | 0.2624 | 10.06 | 0.264 | 12.07 | 0.0518 |
| Hungary | 9.75 | 143 | 272 | 42.3 | 21.03 | 0.0586 | 1.07 | 0.0766 |  |  |
| Luxemburg | 8.65 | 325 | 237 | 39.3 | 16.03 | 0.4841 | 1.06 | 0.0271 |  |  |
| Malta | 19.2 | 1 | 1567 | 41.8 | 23.03 | 0.0712 | 19.04 | 0.0536 | 25.07 | 0.3065 |
| Moldova | 9.45 | 139 | 79 | 36.7 | 22.03 | 0.1716 | 31.04 | 0.0217 |  |  |
| N Macedonia | 9.8 | 741 | 81 | 37.9 | 21.03 | 0.0858 | 3.05 | 0.028 | 26.05 | 0.0808 |
| Netherlands | 9.25 | 30 | 421 | 42.6 | 05.03 | 0.2485 | 7.07 | 0.0928 |  |  |
| Norway | 1.5 | 460 | 17 | 39.2 | 09.03 | 0.2716 | 19.07 | 0.1725 |  |  |
| Poland | 7.85 | 173 | 123 | 40.7 | 14.03 | 0.1562 | 5.04 | 0.0094 | 3.07 | 0.0245 |
| Romania | 8.8 | 414 | 81 | 41.1 | 14.03 | 0.1596 | 31.05 | 0.0498 |  |  |
| Serbia | 10.55 | 473 | 89 | 42.6 | 19.03 | 0.1919 | 1.06 | 0.2547 |  |  |
| Slovenia | 8.9 | 492 | 266 | 44.5 | 17.03 | 0.1301 | 12.06 | 0.0988 |  |  |
| Spain | 13.3 | 660 | 93 | 42.7 | 25.02 | 0.335 | 29.06 | 0.0846 |  |  |
| Sweden | 2.1 | 320 | 23 | 41.2 | 05.03 | 0.2572 | 24.05 | 0.0768 |  |  |
| Switzerland | 5.5 | 1350 | 208 | 42.4 | 04.03 | 0.2388 | 8.06 | 0.0664 |  |  |
| UK | 8.45 | 162 | 280 | 40.5 | 04.03 | 0.2223 | 2.07 | 0.0416 |  |  |
| Ukraine | 8.3 | 175 | 70 | 40.6 | 24.03 | 0.1615 | 25.05 | 0.048 | 6.07 | 0.0165 |
| North America |  |  |  |  |  |  |  |  |  |  |
| Canada | -5.35 | 487 | 4 | 42.2 | 10.03 | 0.2432 | 1.07 | 0.0372 |  |  |
| Cuba | 25.2 | 108 | 102 | 41.5 | 27.03 | 0.0706 | 17.05 | 0.0517 | 24.06 | 0.0741 |
| Domin. Rep. | 24.55 | 424 | 560 | 28.1 | 20.03 | 0.1403 | 1.06 | 0.0305 |  |  |
| El Salvador | 24.45 | 442 | 319 | 27.1 | 10.04 | 0.0783 | 21.04 | 0.0535 | 1.06 | 0.0453 |
| USA | 8.55 | 760 | 34 | 38.1 | 02.03 | 0.2882 | 7.06 | 0.0411 |  |  |
| South America |  |  |  |  |  |  |  |  |  |  |
| Peru | 19.6 | 1555 | 25 | 28 | 25.03 | 0.0915 | 30.6 | 0.0119 |  |  |

For the two variables extracted for third waves, slope and onset date, we found three statistically significant correlations (two tailed P < 0.05). Time since third wave onset correlates negatively with mean altitude (r = −0.50872, two tailed P = 0.031) and positively with country GDP per capita (r = 0.521, two tailed P = 0.0266). The latter correlation could suggest that populations with more income can longer limit interactions, delaying new waves. Time since first wave onset also increases with GDP per capita (r = 0.692, two tailed P > 0.00001). This positive correlation with GDP per capita is not observed for the second wave. Hence, associations between GDP per capita and wave onsets has no simple overarching socio-economic explanation. In addition, these correlations with GDP were not observed for the USA.

The only association that was observed in each dataset, that of countries at large and states of the USA, is the positive association between third wave slope and altitude (for countries, excluding Malta because of its outlier high slope, r = 0.557, two tailed P = 0.016; pooling third wave slopes from both datasets, r = 0.5947, two tailed P = 0.00033, Figure 3). The Malta exception could be due to the highest immigration rate in Europe (about 6% in 2018, after the web site https://ec.europa.eu/eurostat/statisticsexplained/).

**Figure 3.**
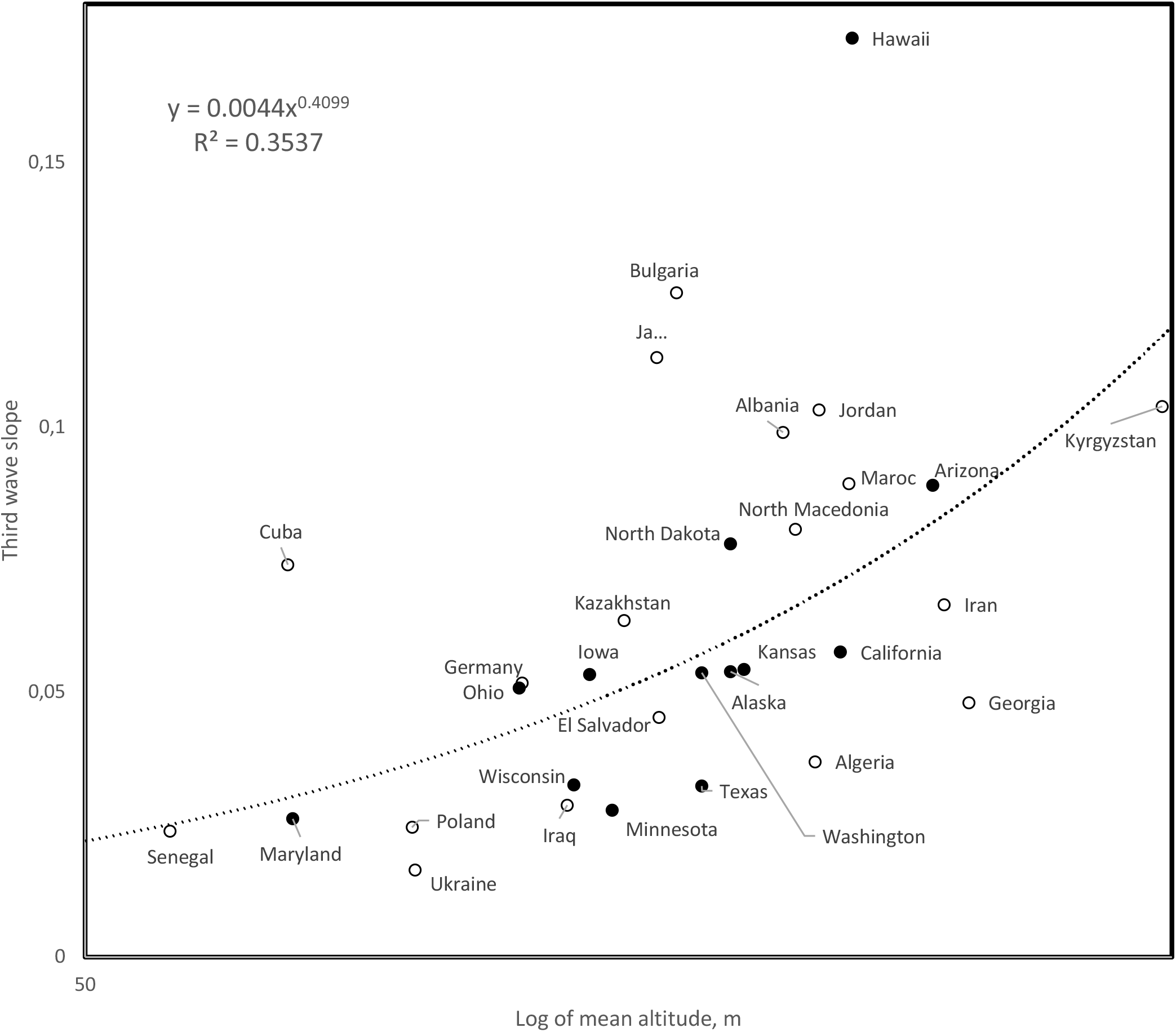
Third wave slopes as a function of mean altitude for 19 countries and 12 states of the USA (open and filled circles, respectively). Correlation calculated after excluding the datapoint for Malta, whose third wave slope is an unexplained outlier (slope = 0.3065, mean altitude at sea level, not included in Figure 3).

## Discussion

### Environmental patterns

The principle of inversion of trends over time, such as in negative heritabilities and for first vs second COVID-19 waves, is partially upheld for third waves of the COVID-19 pandemy in the summer of 2020. Mean country altitude above 900m correlates negatively with first wave slopes, but positively with third wave slopes. There was no pattern with altitude for second wave slopes. Below 900m, first wave slopes increase with altitude, so the change for third wave slopes could reflect adaptation for high spread above 900m. Hypothetically, third wave viruses resist better high elevation UVs. Altitude correlates negatively with temperature, but patterns with altitude are independent of the reported negative correlation between first wave slopes and mean annual temperature (Demongeot et al 2020, Seligmann et al 2020).

Note that third wave slopes are uncorrelated to mean annual temperature, and median age, unlike observations for first and second wave slopes (Demongeot et al 2020, Seligmann et al 2020). They do not correlate with population density, unlike first and second waves in the USA (see above section).

### Intrinsic viral evolution

First/second wave slopes decrease/increase with time since wave onset (Seligmann et al 2020). This could be interpreted as secondary effects due to temperature, because time since wave onset corresponds to temperature increase in countries from the northern hemisphere. This confounding effect of increasing seasonal temperatures is unlikely, because data for Chile and Argentina, southern hemisphere countries with seasonal climates, fit the same patterns with mean annual temperature and time since wave onset as other countries (Seligmann et al 2020).

This suggests that evolution of the viral spread parameter, as estimated by exponential slopes, reflects intrinsic constraints on viral evolution, possibly a deterministic trajectory of serial mutations, such as suggested by parallel evolution of independent SARS-COVID-19 virus populations (Forster et al 2020). Figure 4 plots first, second and third wave slopes as a function of time since February 15, 2020. With few exceptions, first/second wave slopes fit the negative/positive trends reported previously (Seligmann et al 2020). Third wave slopes continue the pattern of increase with time observed for second waves. Notably, the five southern hemisphere countries with seasonal climates (south of the tropic of the Capricorn: Argentina, Chile, South Africa, Australia and New Zealand (the three latter were added to the sample for the sake of this analysis) fit in the overall trends. Hence, slopes for countries transiting from hot to cold climate in spring fit patterns observed for countries transiting at that period from cold to hot seasons.

**Figure 4.**
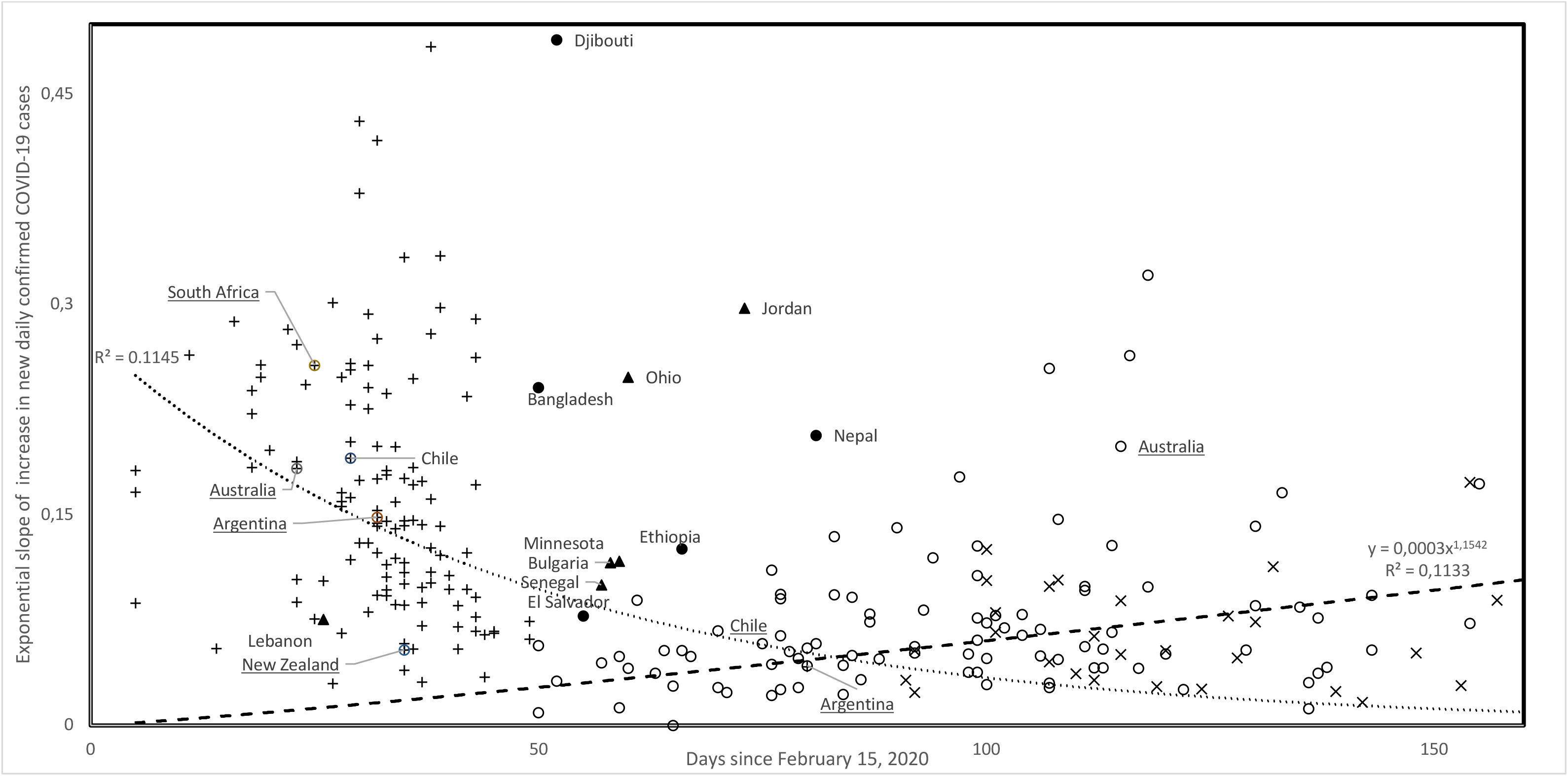
Slopes of first (+), second (O) and third (x) waves as a function of time since February 15, 2020. Filled circles: first wave slopes matching second wave trend; filled triangles: third wave slopes not matching third wave trend. Countries south of the tropic of Capricorn are underlined and fit into overall trends.

Note that for three among five countries with first waves starting after 50 days since February 15 (after April 4), slopes are higher than expected by the overall trend (El Salvador, Ethiopia and Nepal). Within the sample of 123 countries we examine, we found two additional countries/regions with first wave onset after April 4 not included in previous analyses, Bangladesh (4.04, 0.2409) and Djibouti (4.06, 0.489). Both fit this pattern of high first wave slopes that were characteristic of countries with early (February/March) first wave onset dates.

These observations show that a large part of the variation in slopes during the pandemy is due to factors intrinsic to evolutionary trajectories of the virus. This does not exclude environmental effects due to population age structure, density, temperature and altitude, among others. However, inversions of trends with these cofactors at different periods of the pandemy suggest that unknown intrinsic factors have major effects on the evolution of the pandemy.

### Change in life history and trend inversions

Inversion of trends, such as for negative heritabilities, presumably result from drastic environmental changes. For example, levels of channeling of *Sorghum bicolor* plant populations towards developmental trajectories better adapted to NaCl salinity increased with population variability (Seligmann and Amzallag 1995) in the first generation exposed to salinity, but decreased with populational variability in their offspring (Seligmann 1997, 1998). This is in line with the concept that COVID-19 is adapting to its new human host and to various environments inhabited by that host.

Our approach is limited to estimating spread rates during monotonous increases in new daily cases. These might occur at specific periods, possibly when confinement policies are relaxed. It is possible that the hypothesized intrinsic changes in the virus that alter spread parameters occur independently of new case outbreaks, hidden to our method by the lack of outbreak. Alternatively, spread rate changes might be specifically associated with the fast increases in virus populations associated with new case outbreaks. Other analytical methods for estimating spread rates are required to test for these different scenarios.

### Uncertain predictions

The inversion of patterns with time and other variables (altitude, temperature, median population age) for slopes of pandemy waves renders predictions particularly dubious. In addition, the negative correlation between second wave slopes and USA density (Figure 2B) contradicts an accepted epidemiological principle. We see two scenarios for future evolution of COVID-19 spread parameters. Scenario A assumes that by autumn, patterns will get inverted again. Scenario B extrapolates the regression for second and third wave slopes in Figure 4. This predicts that by the end of 2020, slopes would be 0.23 (on average, first, second and third wave slopes are 0.1668, 0.0776 and 0.0731, respectively). Along the same method, slopes will reach the level of mean first wave slopes somewhen in October. A combination of scenarios A and B is also possible, where patterns get inverted at the beginning of the year 2021. Note that scenario C, where slopes reach 0, seems very unlikely.

## Conclusions

1. In the USA first and second wave slopes are not correlated with temperature, median age and time since wave onset, but with population density. The principle of inverted trends between first and second waves upholds, density effects are positive/negative for first/second wave slopes.
2. Third wave slopes (in 13 states of USA and 19 countries) correlate only with altitude, suggesting adaptation for higher spread above 900m. This also inverts a negative trend with altitudes above 900m for first wave slopes.
3. Evolution of viral spread apparently follows intrinsic constraints, independent of environmental factors such as population age structure, density, temperature and altitude. The inversion of correlations between spread parameters and these factors, at different periods (winter, spring, summer), suggests that unknown constraints intrinsic to the virus preponderantly determine pandemic patterns.

## Data Availability

All used data are coming from public data bases explicitely cited

